# Prevalence, risk factors, nature, and nutritional impact of sexual abuse among young girls: A school-based study

**DOI:** 10.64898/2026.03.17.26348669

**Authors:** Niraz Yadav, Aakriti Yadav, Neeru Yadav

## Abstract

Sexual abuse among adolescent girls is underreported in low and middle income countries including Nepal. This study aimed to estimate the prevalence of SA among school girls, examined associated sociodemographic and contextual factors describe the nature and reporting patterns of abuse and assess the relationship with nutritional status. A school based cross sectional study was conducted, among 330 female students (ages 14-19) were selected through simple random sampling from two schools. Data were collected using a validated self-administered questionnaire covering demographic characteristics, abuse experiences, psychosocial responses and reporting patterns. Anthropometric measurements were used to assess BMI-for-age and height-for-age Z scores calculated using WHO AnthroPlus. Logistic regression analysis was used to identify factors independently associated with sexual abuse and adjusted odds ratio with 95% confidence intervals were calculated.

SA prevalence was 33.3%. Most perpetrators were male (61.5%) and known to the victim, 63.3% involved perpetrator use. Reporting was low (16.5%) due to fear (42.2%) and shame (22%). Significant predictors included lower maternal education (AOR=3.03) and living in joint families (AOR=2.34).After adjusting for confounders, SA was strongly associated with thinness (AOR=5.59; 95% CI; 2.54-12.26), severe thinness (AOR=18.81; 95% CI: 4.21-84.07) and stunting (AOR=3.79; 95% CI: 1.88-7.62). One in three girls experienced sexual abuse, which is strongly correlated with growth impairment and malnutrition. These findings suggest that anthropometric deficits may serve as clinical red flags for underlying trauma. Strengthening school-based nursing programs and primary care screening is essential for early identification and safeguarding.

## 1. Introduction

The World Health Organization (WHO) estimates that approximately one in five young girls globally experience sexual abuse before reaching the age of 18. This disproportionately concentrated in low and middle income countries (LMICs) where structural gender inequality, pervasive social stigma and fragmented child-protection systems exacerbate vulnerability [1, 2]. Adolescence represents a particularly vulnerable developmental period, during which frequent exposure to sexual abuse encompassing both contact and non-contact forms can profoundly disrupt educational attainment, mental health, and nutritional status. Such trauma often leads to adverse long term health trajectories and persists as a significant barrier to global public health equity.

In South Asia, including Nepal, sexual violence against adolescent girls remains chronically underreported and under-researched, particularly within school-going populations. Sexual abuse incidence has been taking place across diverse settings, including domestic environments, neighborhoods, schools, streets, workplaces. However deep-seated cultural taboos surrounding sexuality, fear of social exclusion and limited awareness of legal protections contribute to significant underestimation of its true prevalence [3, 4]. Current data suggest that perpetrators are frequently known to the victim and that incidents often occur in institutional or public spaces. Yet, formal reporting to law enforcement is uncommon [5]. As a result, many survivors rarely enter legal or social welfare pathways, often only engaging with the healthcare system indirectly.

From a clinical perspective, adolescents experienced sexual abuse frequently present to primary care clinics or emergency departments with non-specific complaints or psychosomatic complains such as abdominal pain, unexplained injuries, unwanted pregnancy, school absenteeism or nutritional deficits rather than explicit disclosure of trauma [6, 7]. Despite these clinical observations, empirical evidence linking sexual abuse to measurable physical markers such as malnutrition and growth impairment remains sparse particularly in school based studies from LMIC settings. Furthermore, there are critical knowledge gap regarding the specific etiology of abuse, perpetrator-victim relationship and the coping mechanisms with the after-effects of survivor.

This study aimed to estimate the prevalence of sexual abuse among higher secondary school girls in Nepal and identify associated sociodemographic factors. Specifically, we sought to characterize the nature of abuse, evaluate reporting patterns and assess the relationship between a history of abuse and current nutritional status. By integrating epidemiological, psychosocial and anthropometric data this study provides evidence to support school-based health programs, primary care screening, advocating for a holistic and proactive approach to adolescent safeguarding.

## Materials and Methods

### 2.1 Study design and setting

A school-based quantitative cross-sectional study was designed to explore the prevalence and nutritional status on sexually abused young girls. The study was carried out in two randomly selected higher secondary schools of Kathmandu city, Nepal, comprising 15 classrooms. The study area represents a diverse population with varying ages and socioeconomic backgrounds.

### 2.2 Study population

The study population consisted of female students enrolled in grades 11 and 12, aged 14 to 19 years, as this age group is transitional and vulnerable to sexual abuse. A multi-stage sampling technique was employed. First two schools were selected from a comprehensive list of higher secondary institutions using a lottery system. Subsequently, simple random sampling was used to select participants within these schools. Students who did not provide assent or whose Legally Authorized Representatives (LAR) did not provide consent were excluded and participation was voluntary with no incentives offered.

### 2.3 Sample size and sampling method

Data collection occurred from February 1 to March 12, 2024, with a total of 330 students participating. The sample size was calculated from the Cochran formula(n) = z²pq/d², assuming the prevalence (p) of sexual abuse as 28.5% from a previous study conducted [8], at statistics corresponding to the level of 95% confidence interval (z), 5% tolerable error (d) and with estimated 10% non-response rate (q).According to Nepal law, parental consent is required for participants under 16 years of age, and 4% of participants either did not provide consent or were unable to complete the form.

### 2.4 Data collection tools and validity

A structured proforma was developed following extensive literature review. Content validity was assessed by a panel of six experts, including public health officers and general practitioners. The instrument’s validity was quantified using the Item-Level Content Validity Index (I-CVI) and Scale-Level Content Validity Index (S-CVI/Ave) [9]. Items with I-CVI<0.78 were revised or removed. The final instrument demonstrated excellent content validity (S-CVI/Ave= 0.91). The tool was pretested on a small group of 30 students to ensure the participants’ comfort, comprehension and clarity. Feedback from pilot testing was used to refine question wording, sequencing and probing techniques.

The questionnaire was available in both English and Nepali and participants were allowed to choose their preferred language. It consisted of four sections: (1) socio-demographic characteristics, (2) Nature and context of sexual abuse, (3) Psychosocial and reporting responses and (4) Nutritional status parameters.

### 2.5 Data collection procedure

A 20-minute educational session was conducted using PowerPoint slides. We also provided awareness on sexual abuse, responses to sexual harassment and understanding child protection laws (Child Act 2018). We highlight the importance of calling toll-free numbers for help, such as Nepal Police (100), Child Helpline (1098) and the National Women Commission (1145). An awareness video, “KOMAL,” by Childline India Foundation, which was translated into Nepali was also screened [10].

After the class, we emphasized voluntary participation and the right to skip questions or withdraw at any time. The questionnaires were distributed to 345 students along with informed consent, of which 330 returned the following day with signatures from themselves and their parents. Trained public health officers and doctors, assisted in measuring parameter for height, weight measurement completing the forms, provided support and encouraged reporting to trusted individuals or authorities. Participants were thanked for their time and assured of confidentiality about data security. Participation had no incentives offered.

### 2.6 Anthropometric Measurements

Following the questionnaire physical measurements were taken by trained researchers. Height was measured to the nearest 0.1cm using a portable stadiometer and weight was measured to the nearest 0.1kg using a calibrated digital scale. BMI-for-age Z-score (BAZ) and Height-for-age Z-scores (HAZ) were calculated using the WHO AnthroPlus software (version 1.0.4). According to WHO 2007 growth references for adolescents, the following parameters were defined; thinness as BAZ < - 2SD, severe thinness as BAZ < - 3SD, overweight as BAZ > + 1SD, obesity as BAZ > + 2SD and stunting as HAZ < - 2SD[11].

### 2.7 Statistical analysis

Data were entered in EpiData version 3.1. After confirming completeness, the data were exported to IBM SPSS Version 23 for further analysis. Univariate analysis was performed using frequencies, percentages, means, and standard deviations. Multicollinearity among independent variables was assessed with variance inflation factor (VIF) and all variables had VIF values less than 5, indicating no significant multicollinearity [12]. The goodness of fit of the logistic regression model was evaluated by applying Hosmer and Lemeshow chi-square test. Those variables significantly associated in the univariate analysis at 95% confidence level (P- value < 0.01) were included in the multivariate logistic regression model. Adjusted odds ratios (AOR) and Crude odds ratio (COR) with along with 95% confidence intervals were computed to determine the strength of association.

### 2.8 Ethics consideration and confidentiality

Administrative approval was obtained from the chief of the higher secondary school, and ethical approval was granted by the Ethical Review Board of the Nepal Health Research Council (Approval No: 1088). Following NHRC guidelines for research involving minors (aged less than 18years) a dual consent process was utilized. Written informed consent was obtained from the participants’ legally authorized representatives (LAR). Additionally written informed assent was obtained from the participants themselves using a simplified proforma. To protect participant privacy and prevent LAR interference in sensitive disclosures while the LAR consent was obtained at home, the participant assent and data collection were conducted independently in a private school setting. All data were anonymized using participant ID codes and students were informed of their right to withdraw at any stage without penalty.

## 3. Result

### 3.1 Sociodemographic characteristics

A total of 330 adolescent girls participated in the study. Out of which 109 (33.3%) reported experiencing sexual abuse. The mean (+SD) age was 16.64 ± 1.5years ranging from 14 years to 19 years. The majority of participants were in Grade 11 (54.5%) and resided in nuclear families (65.2%). Bivariate analysis (Table 1) revealed that the prevalence of sexual abuse was significantly higher among participants from joint families compared to nuclear families (44.4% vs 27%: p=0.002). Maternal factors were significantly associated with sexual abuse. Higher prevalence was observed among girls whose mothers had at least a secondary education or above (40.5% vs 22%: p=0.001) and among those whose mothers were unpaid workers compared with paid workers (42.5% vs 26.5%; p=0.003). no significant differences in prevalence were found regarding age group, grade or paternal occupation (p=0.01) as shown in Table 1.

**Table 1.** Sociodemographic characteristics of adolescent girls and their Association with Sexual Abuse (n = 330)

| Variables | Total participants (330) |  | Participants being sexually abused |  |  |  | P-value |
| --- | --- | --- | --- | --- | --- | --- | --- |
|  |  |  | Yes (109) |  | No (221) |  |  |
|  | N | % | N | % | N | % |  |
| **Age group(years)** |  |  |  |  |  |  | 0.5 |
| 14 to 16 | 176 | 53.3% | 61 | 34.6% | 115 | 65.4% |  |
| 17 to 19 | 154 | 46.7% | 48 | 31% | 106 | 69% |  |
| **Grade** |  |  |  |  |  |  | 0.12 |
| 11 | 180 | 54.5% | 66 | 36.6% | 114 | 63.4% |  |
| 12 | 150 | 45.5% | 43 | 29% | 107 | 71% |  |
| **GPA Score** |  |  |  |  |  |  |  |
| A | 87 | 26.4% | 24 | 27.6% | 63 | 72.4% | 0.05 |
| B | 43 | 13% | 13 | 30.2% | 30 | 69.8% |  |
| C | 124 | 37.6% | 39 | 31.5% | 85 | 68.5% |  |
| D | 51 | 15.5% | 26 | 51% | 25 | 49% |  |
| No Grade (Ng) | 25 | 7.6% | 7 | 28% | 18 | 72% |  |
| **Family type** |  |  |  |  |  |  | 0.002* |
| Nuclear | 215 | 65.2% | 58 | 27% | 157 | 73% |  |
| Joint | 115 | 34.8% | 51 | 44.4% | 64 | 55.6% |  |
| **Father's education** |  |  |  |  |  |  | 0.415 |
| Primary and below | 79 | 24% | 23 | 29% | 56 | 71% |  |
| Secondary and above | 251 | 76% | 86 | 34.2% | 165 | 65.8% |  |
| **Mother's education** |  |  |  |  |  |  | 0.001* |
| primary and below | 135 | 41% | 30 | 22% | 105 | 78% |  |
| secondary and above | 195 | 59% | 79 | 40.5% | 116 | 59.5% |  |
| **Father's Occupation** |  |  |  |  |  |  | 0.9 |
| Paid | 296 | 90% | 98 | 33% | 198 | 67% |  |
| Unpaid | 34 | 10% | 11 | 32.3% | 23 | 67.7% |  |
| **Mother's Occupation** |  |  |  |  |  |  | 0.003* |
| Paid | 196 | 59.4% | 52 | 26.5% | 144 | 73.5% |  |
| Unpaid | 134 | 40.6% | 57 | 42.5% | 77 | 57.5% |  |
| **Knowledge on sexual abuse topic** |  |  |  |  |  |  | 0.00* |
| Do have | 132 | 40% | 28 | 21% | 104 | 79% |  |
| Don't have | 198 | 60% | 81 | 41% | 117 | 59% |  |

Note: \*P- value 0.01 is significant

### 3.2 Prevalence and nature of Sexual Abuse

Among these victims, perpetrators were predominantly male (61.5%) and older than victim (60%). Strangers (39.4%) or friends (31.2%) were most common perpetrators, followed by family members (15.6%) and teachers (13.8%). Substance use by the perpetrator was noted in 63.3% of incidents. Regarding the circumstances, More than half of the incidents occurred within the 12 months (58.7%) preceding the study. Abuse most frequently took place on the street (31.4%), clubs/restaurants (30.3%), schools (22.9%) and homes (15.6%). Verbal abuse (39.4%) and inappropriate kissing or hugging (29.4%) were the most commonly reported forms of abuse as shown in Table 2.

**Table 2.** Nature and circumstances of a victims (n=109)

| **Variables** | **Category** | **N(109)** | **%** |
| --- | --- | --- | --- |
| **Sex of perpetrator** |  |  |  |
|  | Male | 67 | 61.5% |
|  | Female | 33 | 30.2% |
|  | Both | 9 | 8.3% |
| **Age of perpetrator** |  |  |  |
|  | Elder than victim | 65 | 60% |
|  | Equal age | 37 | 34% |
|  | Younger | 7 | 6% |
| **Relationship with perpetrator** |  |  |  |
|  | Stranger | 43 | 39.4% |
|  | Friend | 34 | 31.2% |
|  | Family member | 17 | 15.6% |
|  | Teacher | 15 | 13.8% |
| **Motive to do sexual abuse** |  |  |  |
|  | Male domination | 34 | 31.2% |
|  | Drug facilitated sexual violence | 33 | 30.3% |
|  | Short term relationship | 21 | 19.3% |
|  | Sexual gratification and personal enjoyment | 8 | 7.3% |
|  | Distorted thinking | 13 | 11.9% |
| **Consumed any toxic substances** |  |  |  |
|  | Yes | 69 | 63.3% |
|  | No | 40 | 36.7% |
| **Time since Abuse** |  |  |  |
|  | Within past 12months | 64 | 58.7% |
|  | Greater than 12months | 45 | 41.3% |
| **Place of Incident** |  |  |  |
|  | Street | 34 | 31.2% |
|  | Club/Restaurant | 33 | 30.3% |
|  | School/Institute | 25 | 22.9% |
|  | Home | 17 | 15.6% |
| **Nature of abuse** |  |  |  |
|  | Verbally abuse | 43 | 39.4% |
|  | Inappropriate kissing and hugging | 32 | 29.4% |
|  | Exposure to pornography or nudity | 15 | 13.8% |
|  | Physically abuse and touching of genital | 12 | 11% |
|  | Forced to have sex | 7 | 6.4% |

### 3.3 Psychosocial impact and Reporting

The psychosocial burden was substantial. 46% of victims missed school on the following day the incident and 54% reported feelings of self-blame. Parental support was low with only 26.6% of victims perceiving that their parents understood their situation. Consequently, formal reporting was rare as only16.5% of incidents were reported to the police. The main reasons for non-reporting included fear of serious harm from the accused (42.2%) shame or embarrassment (22%), lack of awareness regarding available resources (20.2%) which is shown in Table 3.

**Table 3.**
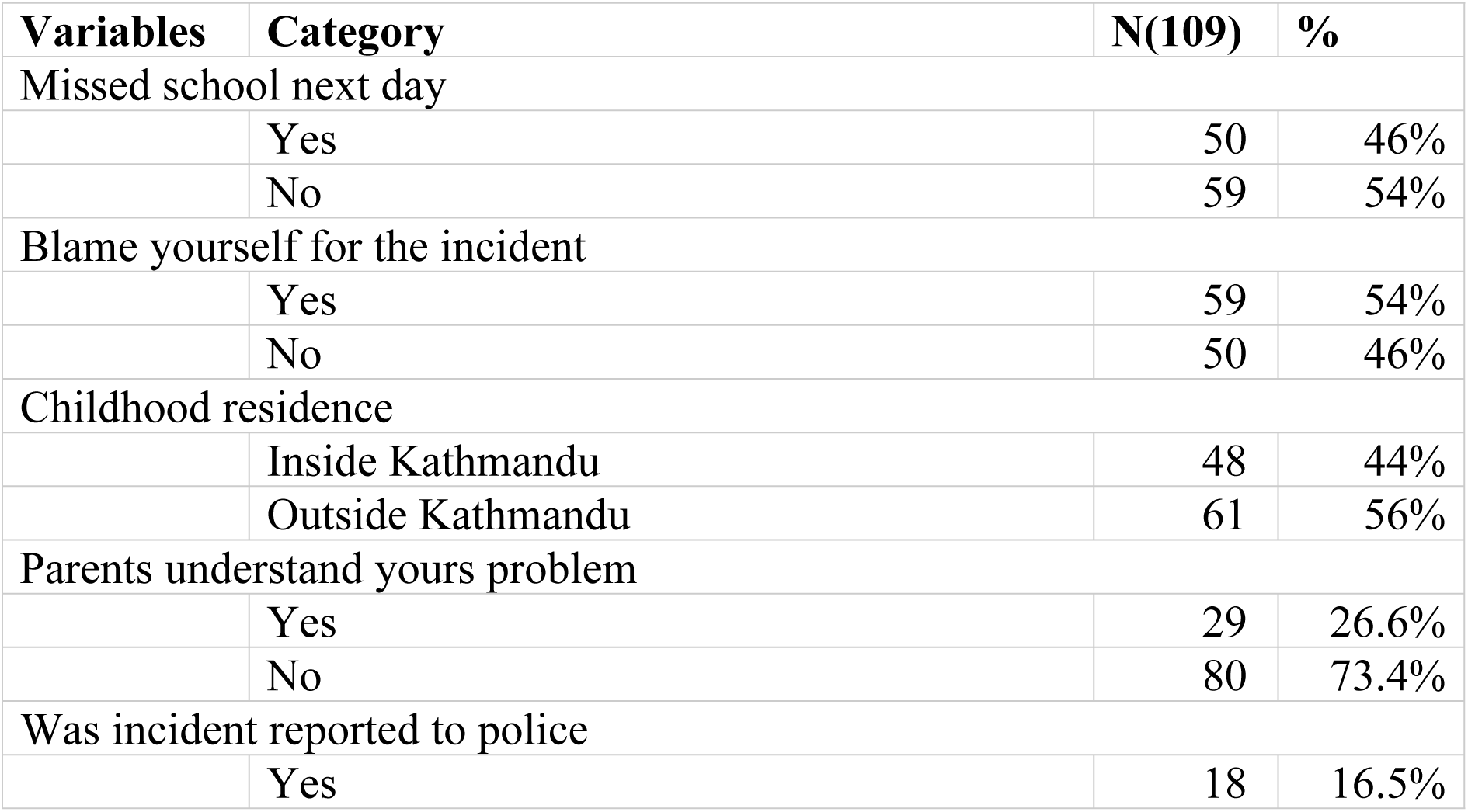
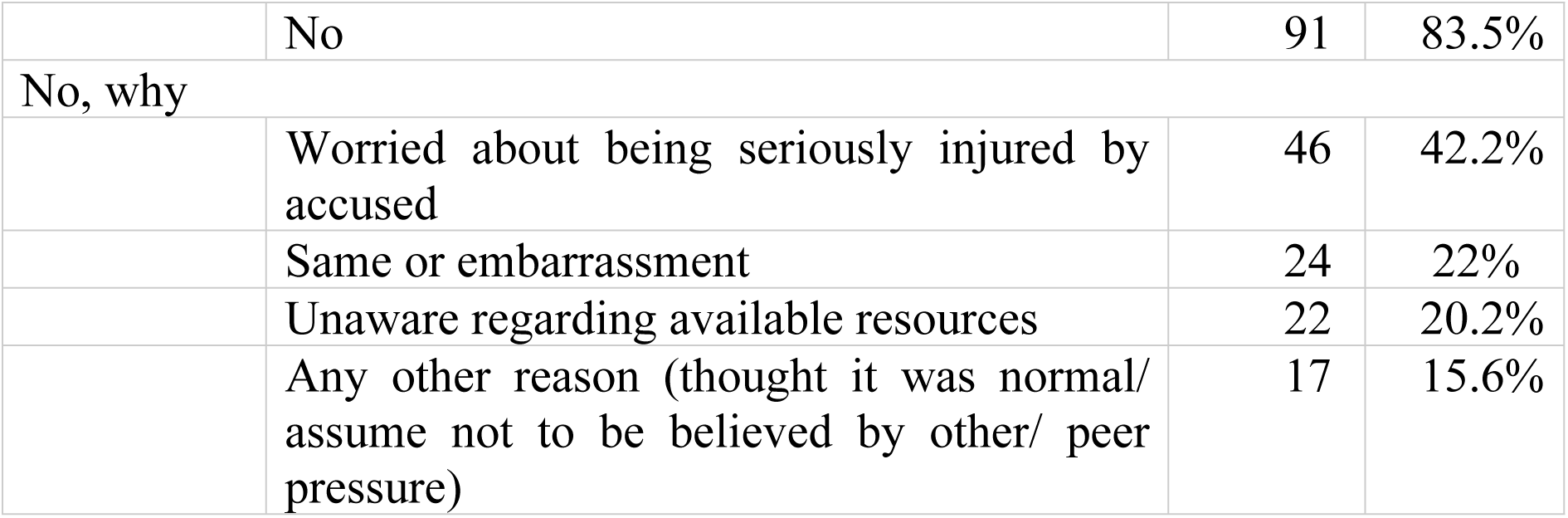
Psychosocial and Reporting Responses of Victims (n = 109)

| Variables | Category | N(109) | % |
| --- | --- | --- | --- |
| Missed school next day |  |  |  |
|  | Yes | 50 | 46% |
|  | No | 59 | 54% |
| Blame yourself for the incident |  |  |  |
|  | Yes | 59 | 54% |
|  | No | 50 | 46% |
| Childhood residence |  |  |  |
|  | Inside Kathmandu | 48 | 44% |
|  | Outside Kathmandu | 61 | 56% |
| Parents understand yours problem |  |  |  |
|  | Yes | 29 | 26.6% |
|  | No | 80 | 73.4% |
| Was incident reported to police |  |  |  |
|  | Yes | 18 | 16.5% |
|  | No | 91 | 83.5% |
| No, why |  |  |  |
|  | Worried about being seriously injured by accused | 46 | 42.2% |
|  | Same or embarrassment | 24 | 22% |
|  | Unaware regarding available resources | 22 | 20.2% |
|  | Any other reason (thought it was normal/ assume not to be believed by other/ peer pressure) | 17 | 15.6% |

### 3.4 Nutritional status as a clinical marker

Nutritional status was significantly associated with a history of sexual abuse (p=0.001). Victims exhibited higher rates of Thinness (42%) and severe thinness (62.5%) were more prevalent among sexually abused girls compared to non-abused peers (Fig 1). Stunting was also higher among victims (51.5%), highlighting malnutrition as a potential clinical marker of abuse (Fig 2).

**Fig 1.**
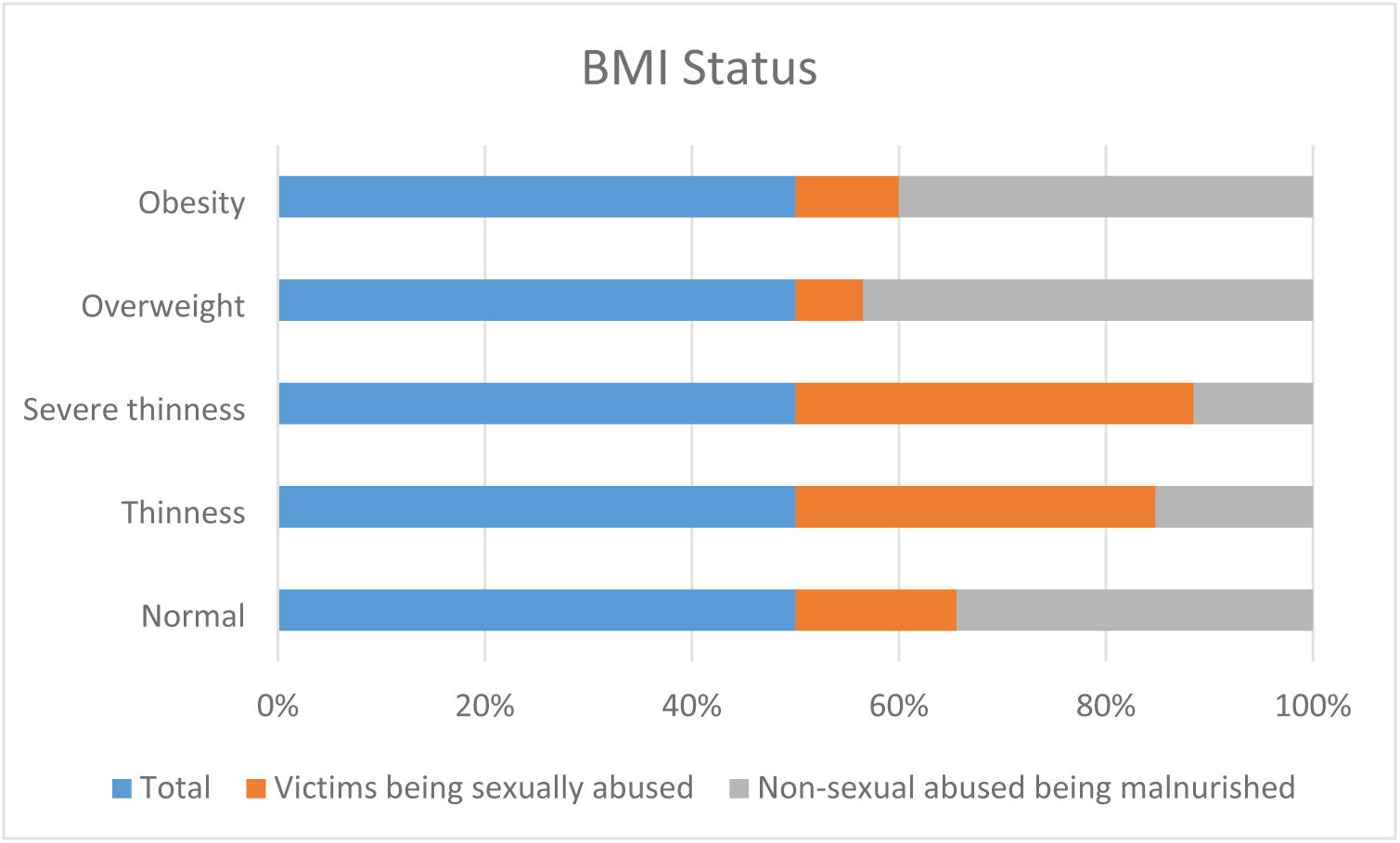
**Participants BMI status**

**Fig 2.**
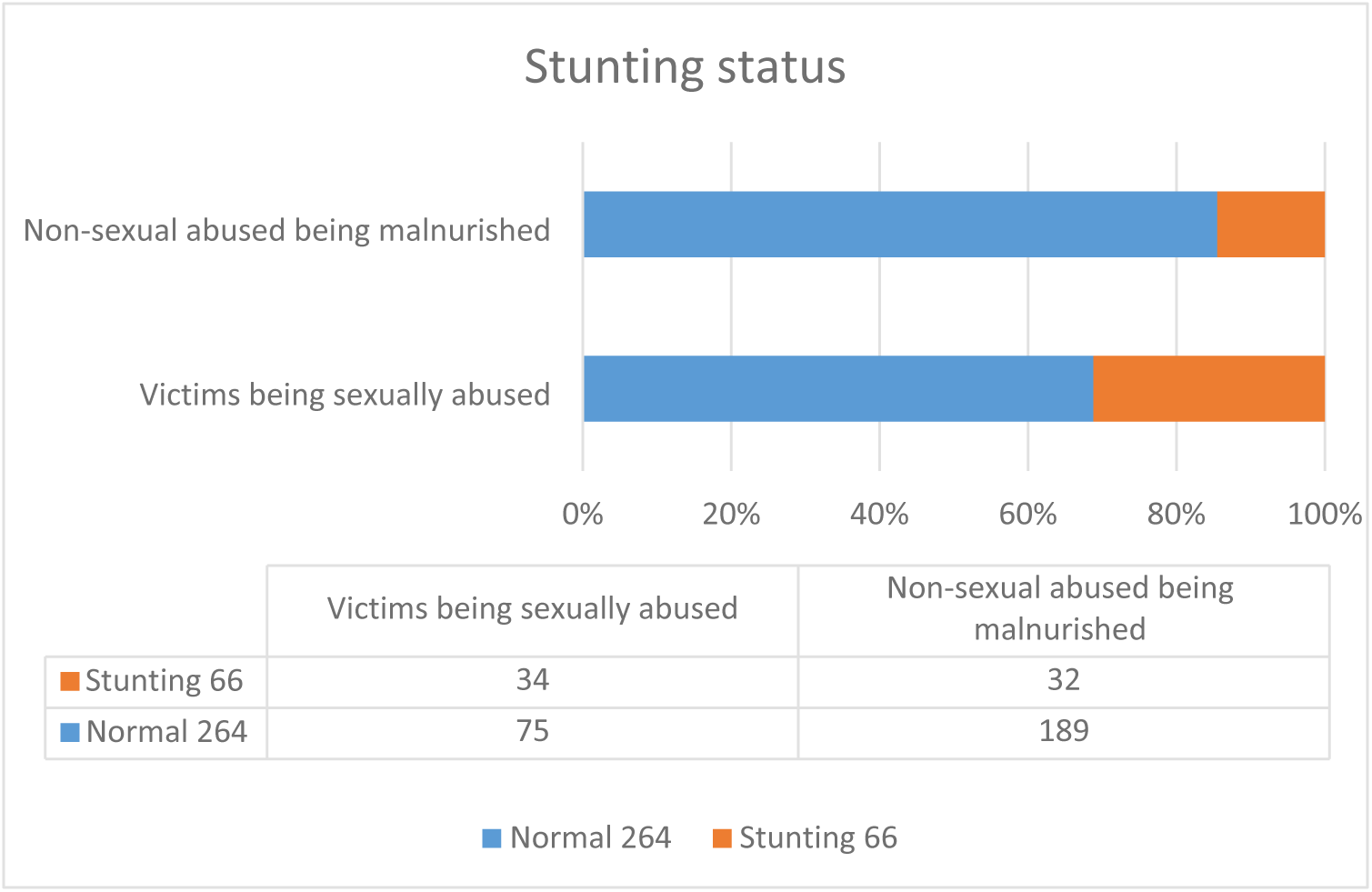
**Participants stunting status**

### 3.5 Factors associated with Sexual Abuse

The multivariate logistic regression model (Table 4) identified several independent predictors of sexual abuse. Girls whose mothers had primary or lower education had a three-fold increase in the odds of abuse (AOR=3.03, 95% CI: 1.63-5.64). Maternal paid occupation (AOR=1.79, 95% CI: 1.02-3.15) and living in nuclear families (AOR=2.34, 95% CI: 1.29-4.15) were also significant predictors. Lack of sexual education showed a strong association (AOR=3.45, 95% CI: 1.83-6.49). The stronger clinical associations were related to anthropometric markers. After adjusting for socioeconomic confounders, thinness (AOR=5.59, 95% CI: 2.54-12, 26), severe thinness (AOR=18.81, 95% CI: 4.21-84.07) were significantly associated with a history of abuse. Also stunting (AOR=3.79, 95% CI: 1.88, 7, 62) significantly increasing risk.

**Table 4.**
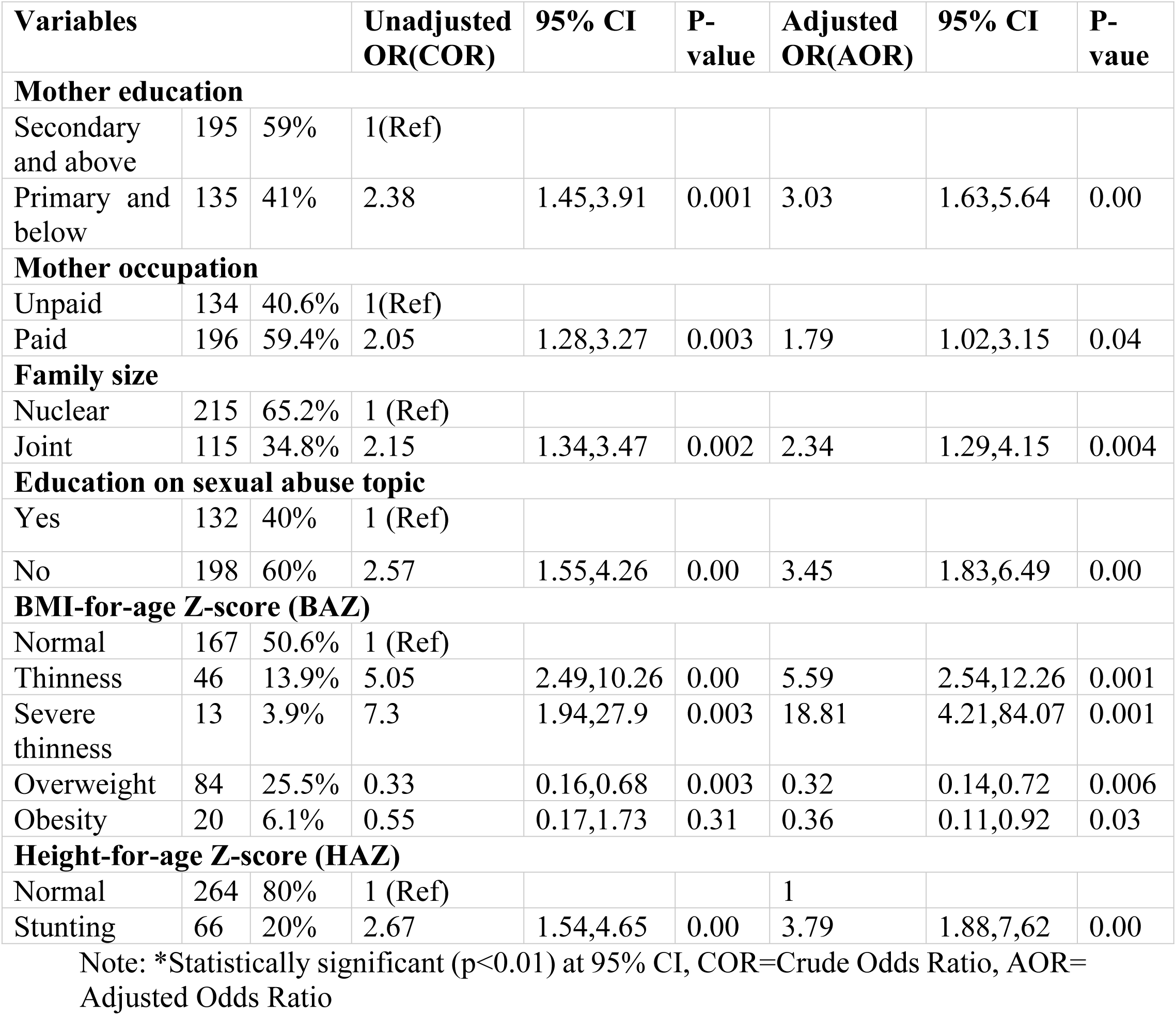
Multivariate factors associated with sexual abuse.

| Variables |  |  | Unadjusted<br>OR(COR) | 95% CI | P-value | Adjusted<br>OR(AOR) | 95% CI | P-vaue |
| --- | --- | --- | --- | --- | --- | --- | --- | --- |
| **Mother education** |  |  |  |  |  |  |  |  |
| Secondary<br>and above | 195 | 59% | 1(Ref) |  |  |  |  |  |
| Primary and<br>below | 135 | 41% | 2.38 | 1.45,3.91 | 0.001 | 3.03 | 1.63,5.64 | 0.00 |
| **Mother occupation** |  |  |  |  |  |  |  |  |
| Unpaid | 134 | 40.6% | 1(Ref) |  |  |  |  |  |
| Paid | 196 | 59.4% | 2.05 | 1.28,3.27 | 0.003 | 1.79 | 1.02,3.15 | 0.04 |
| **Family size** |  |  |  |  |  |  |  |  |
| Nuclear | 215 | 65.2% | 1 (Ref) |  |  |  |  |  |
| Joint | 115 | 34.8% | 2.15 | 1.34,3.47 | 0.002 | 2.34 | 1.29,4.15 | 0.004 |
| **Education on sexual abuse topic** |  |  |  |  |  |  |  |  |
| Yes | 132 | 40% | 1 (Ref) |  |  |  |  |  |
| No | 198 | 60% | 2.57 | 1.55,4.26 | 0.00 | 3.45 | 1.83,6.49 | 0.00 |
| **BMI-for-age Z-score (BAZ)** |  |  |  |  |  |  |  |  |
| Normal | 167 | 50.6% | 1 (Ref) |  |  |  |  |  |
| Thinness | 46 | 13.9% | 5.05 | 2.49,10.26 | 0.00 | 5.59 | 2.54,12.26 | 0.001 |
| Severe<br>thinness | 13 | 3.9% | 7.3 | 1.94,27.9 | 0.003 | 18.81 | 4.21,84.07 | 0.001 |
| Overweight | 84 | 25.5% | 0.33 | 0.16,0.68 | 0.003 | 0.32 | 0.14,0.72 | 0.006 |
| Obesity | 20 | 6.1% | 0.55 | 0.17,1.73 | 0.31 | 0.36 | 0.11,0.92 | 0.03 |
| **Height-for-age Z-score (HAZ)** |  |  |  |  |  |  |  |  |
| Normal | 264 | 80% | 1 (Ref) |  |  | 1 |  |  |
| Stunting | 66 | 20% | 2.67 | 1.54,4.65 | 0.00 | 3.79 | 1.88,7.62 | 0.00 |

Note: \*Statistically significant (p<0.01) at 95% CI, COR=Crude Odds Ratio, AOR=
Adjusted Odds Ratio

## 4. Discussion

This school-based study reveals a high prevalence of sexual abuse among adolescent girls in Kathmandu (33.3%), establishing a significant correlation between such trauma and poor nutritional outcomes. Our findings fall within the global meta-analytic range 8-31% [13], though it is higher than the 11.5% previously reported in Nepal [14] and lower than the 53.2% reported in India [15].

Our analysis identifies distinct socio-demographic risks. Adolescent girls from joint families and those whose mothers had lower educational attainment were at significantly increased risk, consistent with prior south Asian evidence [16]. This aligns with the theory that maternal empowerment and higher educational levels serve as critical protective factors against household and community level vulnerability [16, 17]. Regarding maternal occupation, mothers in paid employment showed a higher risk compared to those in unpaid roles, this likely reflects a gap in protective supervision which may leave young girls more vulnerable to exploitation and less likely to share their experiences due to a lack of immediate parental guidance [16, 17, 18].

The circumstances surrounding the incidents provide critical insights for preventive interventions. Findings reveal that perpetrators were predominantly male, older than the victims and often individuals within the victims’ social periphery such as acquaintances or strangers. A significant proportion occurred in public or semi-public spaces including streets, clubs and educational institutions suggesting a breach in community level safeguarding [16, 17]. Notably, substance use by perpetrators was reported in nearly two thirds of the cases, reinforcing established evidence that alcohol and drug consumption act as primary facilitators of sexual violence [5, 19]. The most prevalent forms of abuse verbal harassment, non-consensual kissing or hugging and forced exposure to pornography points toward a pattern of abuse driven by motives of male domination and short term exploitation [20]. These patterns underscore the urgent need for safe public infrastructure and robust school based protection policies.

A profound challenge identified is the pervasive underreporting of abuse which masks the true scale of the crisis. While some incidents were documented, the majority remained undisclosed due to fear of physical retaliation by the accused, deep seated feelings of shame or embarrassment and a fundamental lack of awareness regarding the nature of the abuse. These individual barriers are exacerbated by social stigma and a documented lack of parental understanding, which often renders formal police reporting rare [13, 21]. The immediate impact of abuse is evidenced by school absenteeism on the next day following an incident which is a clear marker of acute distress and fear if disbelief. Adolescent victims frequently navigate complex internal conflicts including guilt and fear of disbelief which mirror global systemic barriers to disclosure [22]. Such persistent feelings if embarrassment and lack of support serve as primary catalysts for PTSD [23, 24]. These psychological disorders frequently disrupt nutritional habits, consequently our study demonstrates a robust association between sexual abuse and adverse nutritional outcomes. We observed a higher prevalence of thinness, severe thinness and stunting among abused girls. Particularly in the 16-19 age group where anthropometric data (Height, Weight and BMI) are typically stable these findings suggest a link between stunting and chronic trauma rather than transient pubertal changes. This supports that childhood maltreatment and chronic stress may disrupt the physiological and endocrine systems essential for growth and nutrition [25].

In Nepal, existing efforts to address sexual abuse include hospital based services through One-Stop Crisis Management Centers (OCMC) [27], NGO led rehabilitation such as Maiti Nepal [28]. Additionally United Nations Population Fund Nepal implements gender-based violence prevention and response programs [29] while regulatory measures by the Nepal Telecommunications Authority include restricting access to pornographic websites. However since this study denotes 58.7% victims were effected within 12 months which may rise further immediate concern and policy to prevent decrease the prevalence. Furthermore, the lack of difference in abuse rates between girls residing inside and outside Kathmandu suggests that vulnerability is a nationwide issue requiring a unified response. To address this the

Nepal government recently launched the “One School, One Nurse” (OSON) initiative to provide adolescent reproductive health and psychosocial support [30]. However a critical gap remains while government schools are benefited from funding for nursing, private schools currently lack similar regulatory mandates as in our two study sites. Thus further strengthening coordination between such initiatives in between schools and primary care based screening may improve early identification and mitigate both psychosocial and nutritional consequences of abuse.

### 4.1 Limitations and strengths

This study provided school based evidence on the prevalence, nature and risk factors of sexual abuse among adolescent girls in Nepal a setting with limited prior data. It is among the few studies to examine the association between sexual abuse and nutritional outcomes highlighting thinness, severe thinness and stunting as important health correlates. The study used structured data collection and included multiple dimensions sociodemographic, psychosocial and nutritional allowing for a comprehensive understanding of vulnerability and consequences.

The cross-sectional design prevents causal inference. Data relied on self-report which may underestimate prevalence due to stigma, fear, or recall bias. The study was limited to school going adolescents, potentially excluding out of school youth who may experience higher risk.

## 5. Conclusion

Sexual abuse among adolescent girls in Nepal remains a significant and underreported public health concern with one in three of participants reporting experiences of abuse. The study highlights that abuse commonly occurs in everyday social environments and is rarely reported due to fear, stigma and lack of awareness of available support systems. Importantly sexually abused girls demonstrated significantly higher rates of thinness, severe thinness and stunting suggesting that poor nutritional status may serve as a potential clinical indicator of chronic psychosocial stress. As adolescents spend a large portion of their time in school where they develop beliefs, values and social understanding, school environments play a critical role in prevention and awareness. Strengthening school-based education, improving parental engagement and integrating screening within primary healthcare and school health programs are essential to protect vulnerable adolescents.

## Data Availability

The data will be available.

## Acknowledgement

The authors would like to acknowledge the people who directly and indirectly contributed to the study. We would like to express gratitude to the higher secondary schools of Kathmandu for granting permission to conduct the study and facilitating the procedure.

## Author contributions

Conceptualization: Niraz Yadav, Aakriti Yadav

Methodology: Niraz Yadav, Aakriti Yadav, Neeru Yadav

Data Collection: Neeru Yadav, Aakriti Yadav, Niraz Yadav

Formal Analysis: Niraz Yadav, Aakriti Yadav

Investigation: Niraz Yadav, Aakriti Yadav

Writing – Original Draft: Niraz Yadav

Writing – Review & Editing: Niraz Yadav, Neeru Yadav, Aakriti Yadav

Supervision: Niraz Yadav

